# Exhaled CO_2_ as COVID-19 infection risk proxy for different indoor environments and activities

**DOI:** 10.1101/2020.09.09.20191676

**Authors:** Zhe Peng, Jose L. Jimenez

**Affiliations:** Cooperative Institute for Research in Environmental Sciences and Department of Chemistry, University of Colorado, Boulder, Colorado 80309, USA

## Abstract

CO_2_ is co-exhaled with aerosols containing SARS-CoV-2 by COVID-19 infected people and can be used as a proxy of SARS-CoV-2 concentrations indoors. Indoor CO_2_ measurements by low-cost sensors hold promise for mass monitoring of indoor aerosol transmission risk for COVID-19 and other respiratory diseases. We derive analytical expressions of CO_2_-based risk proxies and apply them to various typical indoor environments. The relative infection risk in a given environment scales with excess CO_2_ level, and thus keeping CO_2_ as low as feasible in a space allows optimizing the protection provided by ventilation. We show that the CO_2_ level corresponding to a given absolute infection risk varies by over 2 orders of magnitude for different environments and activities. Although large uncertainties, mainly from virus exhalation rates, are still associated with infection risk estimates, our study provides more specific and practical recommendations for low-cost CO_2_-based indoor infection risk monitoring.

**Table of Contents Graphic:** 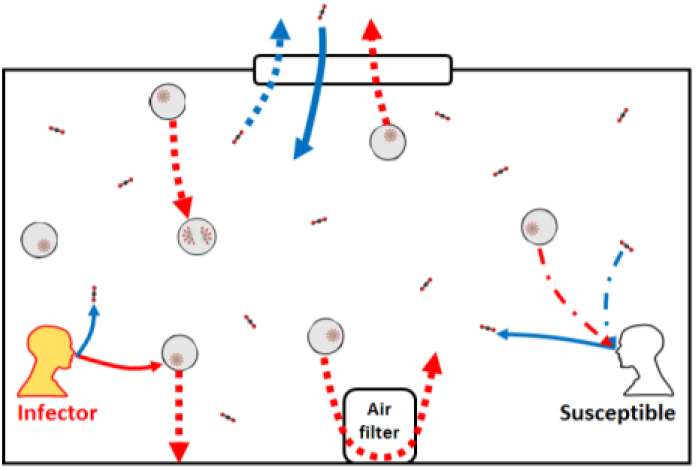

## Introduction

Coronavirus disease 2019 (COVID-19) is currently sweeping the world and causing major losses of human lives.^1^ Lockdowns imposed to various extent worldwide for the COVID-19 transmission reduction are not supposed to be long-term measures, otherwise they would lead to unaffordable social and economic costs. On the other hand, resumption of social, educational, and business activities raises concerns about transmission resurgence.

In last few months, there has been rapidly mounting evidence for COVID-19 transmission via aerosols,^2–5^ i.e., severe acute respiratory syndrome coronavirus 2 (SARS-CoV-2)-containing particles with diameters <100 µm that can float in the air for minutes to hours. Such aerosols have been detected in exhaled air of COVID-19 patients^6^ and in hospital air^7,8^ and the behaviors of smaller ones among them out of proximity of sources have been shown to be similar to gas.^9,10^ Transmission is much easier indoors than outdoors, which is most consistent with aerosols.^4,11,12^ As humans spend most time in indoor environments, where air volumes are limited and virus-laden aerosols may easily accumulate, mitigation of indoor COVID-19 transmissions is a subject of high interest^13,14^ and is key to a successful societal and economic reopening. Practical, affordable, and widely applicable measures to monitor and limit indoor transmission risks are urgently needed.

Direct measurements of virus-containing aerosols are extremely difficult and slow. Indoor CO_2_ has been suggested as an indicator of ventilation of indoor spaces in 19^th^ century,^15^ and more recently as a practical proxy of respiratory infectious disease transmission risk,^16^ as pathogen-containing aerosols and CO_2_ are co-exhaled by those infected (Fig. 1). Since background (ambient) CO_2_ level is almost stable and indoor excess CO_2_ is usually only from human exhalation, measurements of indoor CO_2_ concentration by low-cost CO_2_ sensors can often be good indicators of infection risk and suitable for mass deployment.^17,18^ However, the CO_2_ level corresponding to a given COVID-19 infection risk is largely unknown. A few guideline limit concentrations have been proposed, but without solid and quantitative basis.^19,20^ In particular, only a single CO_2_ threshold was recommended in each of these proposed guidelines. Whether a single CO_2_ concentration ensures low COVID-19 infection risk in all common indoor environments remains an open question, but is also critical for effective CO_2_-based mass risk monitoring.

**Figure 1.**
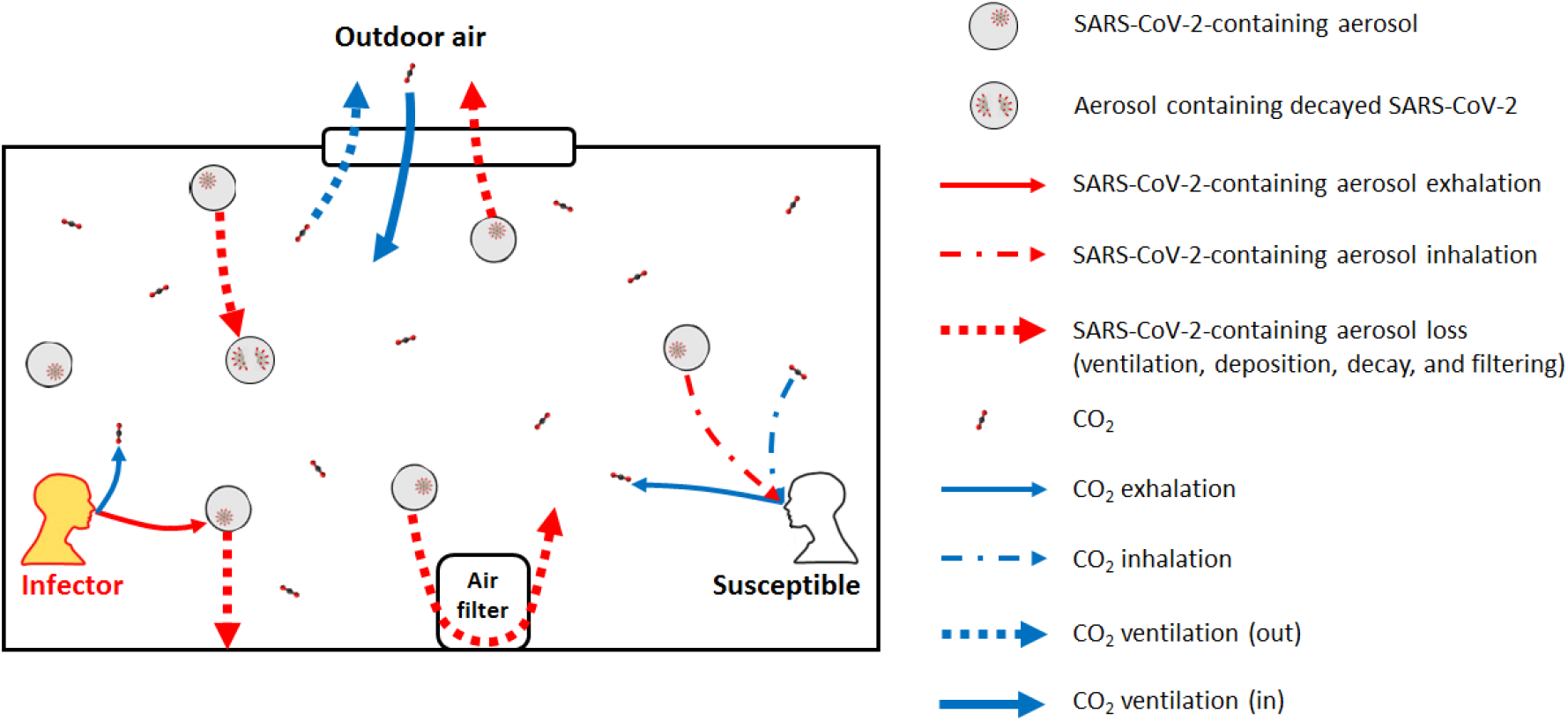
Schematic illustrating the exhalation, inhalation, and other loss processes of SARS-CoV-2-containing aerosols and the exhalation, inhalation, and other source and sink of CO_2_ in an indoor environment.

In this study, we derive the analytical expressions of the probability of indoor COVID-19 infection through room-level aerosol transmission only (i.e., assuming social distance is kept so that close proximity aerosol and droplet pathways are eliminated; fomite transmission is not included), human-exhaled CO_2_ concentration, and subsequently a few CO_2_-based quantities as infection risk proxies. Based on available data, we apply these expressions to common indoor settings to answer the abovementioned open question.

### Derivation of CO_2_-based infection risk proxies

To derive the SARS-CoV-2 aerosol concentration in indoor air, we assume well-mixed air (Fig. 1). The degree of inhomogeneity can be easily quantified with portable low-cost sensors. If significant inhomogeneity in indoor air is present, the indoor space can often be approximated as several compartments, each of them having relatively well-mixed air. Ventilation with outdoor air, virus decay and deposition onto surfaces, and additional control measures (e.g., air filtration and use of germicidal UV) result in losses of infective virus from indoor air. Other sinks (e.g., inhalation by humans and animals indoors) are assumed to be insignificant. This model will underestimate the risk in environments with significant non-respiratory sources of infective aerosols (e.g., bathrooms due to toilet flushing, resuspension in healthcare facilities due to personal-protective-equipment donning/doffing). The amount of the virus infectious doses (*n* – “quanta”) inhaled by a susceptible person determines their probability of infection (*P*) (see Table S1 for the list of symbols in this study). According to the Wells-Riley model of aerosol infection,^21^

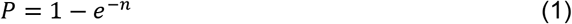

One SARS-CoV-2 quantum corresponds to a probability of infection of 1-1/*e* (63%). The expected value of *n* (⟨*n*⟩) for an originally uninfected person corresponding to a given level of immunity in local population (probability of an occupant being immune, *η*_*im*_), can be calculated as follows

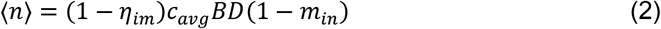

where *c*_*avg*_, *B, D*, and *m*_*in*_ are the average virus concentration (quanta m^-3^), breathing rate of the susceptible person (m^3^ h^-1^), duration of the event (h), and mask filtration efficiency for inhalation, respectively. (1 − *η*_*im*_) is included since quanta inhaled by an immune uninfected individual will not lead to infection and should be excluded. Under the assumption of no occupants and no SARS-CoV-2 in the indoor air at the start of the event, the analytical expression of the expected value of *c*_*avg*_ based on the prevalence of infectors in local population (probability of an occupant being infector, *η*_*I*_), <*c*_*avg*_>, is (see Section S1 for the derivation)

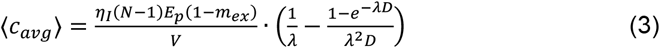

where *N* is number of occupants, *E*_*p*_ is the SARS-CoV-2 exhalation rate by an infector (quanta h^-1^), *m*_*ex*_ mask filtration efficiency for exhalation, *V* indoor environment volume (m^3^), and *λ* first-order overall rate constant of the virus infectivity loss (h^-1^) that includes the ventilation with outdoor air and all other virus removal and deactivation processes.

If there are no other significant CO_2_ sources/sinks (e.g., gas/coal stove and pets/plants), i.e., if indoor excess CO_2_ (relative to the background outdoor level) production is only due to human exhalation and its loss is ventilation, similar quantities for CO_2_ can be expressed as follows (see Section S1 for the derivation)

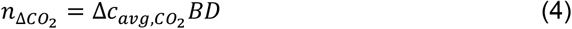

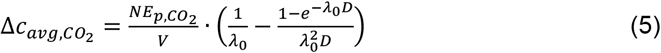

where 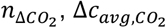, and 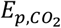 are inhaled excess (human-exhaled) CO_2_ volume (m^3^), excess CO_2_ volume mixing ratio, and CO_2_ exhalation rate per person (m^3^ h^-1^), respectively, and *λ*_0_ is the ventilation rate (h^-1^).

When *P* is low, as it should be for a safe reopening, *P* ≈ *n*. As airborne SARS-CoV-2 and excess CO_2_ are co-exhaled and co-inhaled, in principle 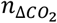 can be a proxy of ⟨*n*⟩, and thus *P*. The ratio of 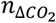 to ⟨*n*⟩ (in m^3^ quantum^-1^),

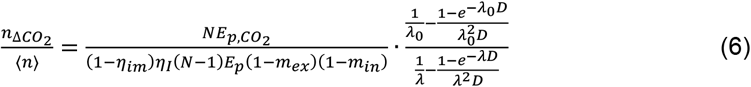

indicates the *volume* of inhaled excess CO_2_ corresponding to a unit inhaled quantum. However, this quantity, involving inhaled CO_2_ volume that is difficult to measure, is not practical for widespread transmission risk monitoring, which usually requires a fast decision-making process simply based on indoor CO_2_ concentration reading (usually in ppm) of a low-cost sensor. Therefore, we propose, as another proxy of the risk of an environment with *η*_*I*_ = 0.1%, reference excess CO_2_ level 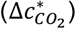, i.e., the volume mixing ratio of excess CO_2_ that an uninfected individual inhales for a typical duration (1 h) in that environment for a typical probability of infection (0.01%).

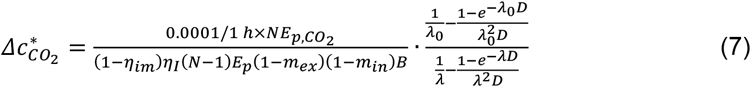

This quantity is closely related to the excess CO_2_ level corresponding to the unity basic reproduction number (*R*_0_)^16^ (see Section S2), and can be directly and easily compared to CO_2_ sensor readings. The ratio of the excess CO_2_ reading to 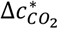 is that of the probability of infection of an originally uninfected person in that environment for 1 h to 0.01%. 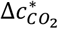 scales (roughly) linearly with most of the parameters in Eq. 7 (see discussions below). *P* = 0.01% being chosen as reference does *not* imply safety at this *P* in all situations, since when *N* and/or *D* are large, and/or the event is repeated many times (e.g., in school/university settings), the overall probability of infection for one susceptible person and/or total infections may still be significant.

## Results and Discussion

Reference excess CO_2_ level is a function of a number of variables. A priori, varying any of them can result in a different value of 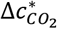 even for similar settings. As an example, we study a set of model cases for a typical university class. The cases are specified in Table S2. The 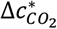 and 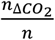 in these cases are shown in Figs. 2A and S1A, respectively.

**Figure 2.**
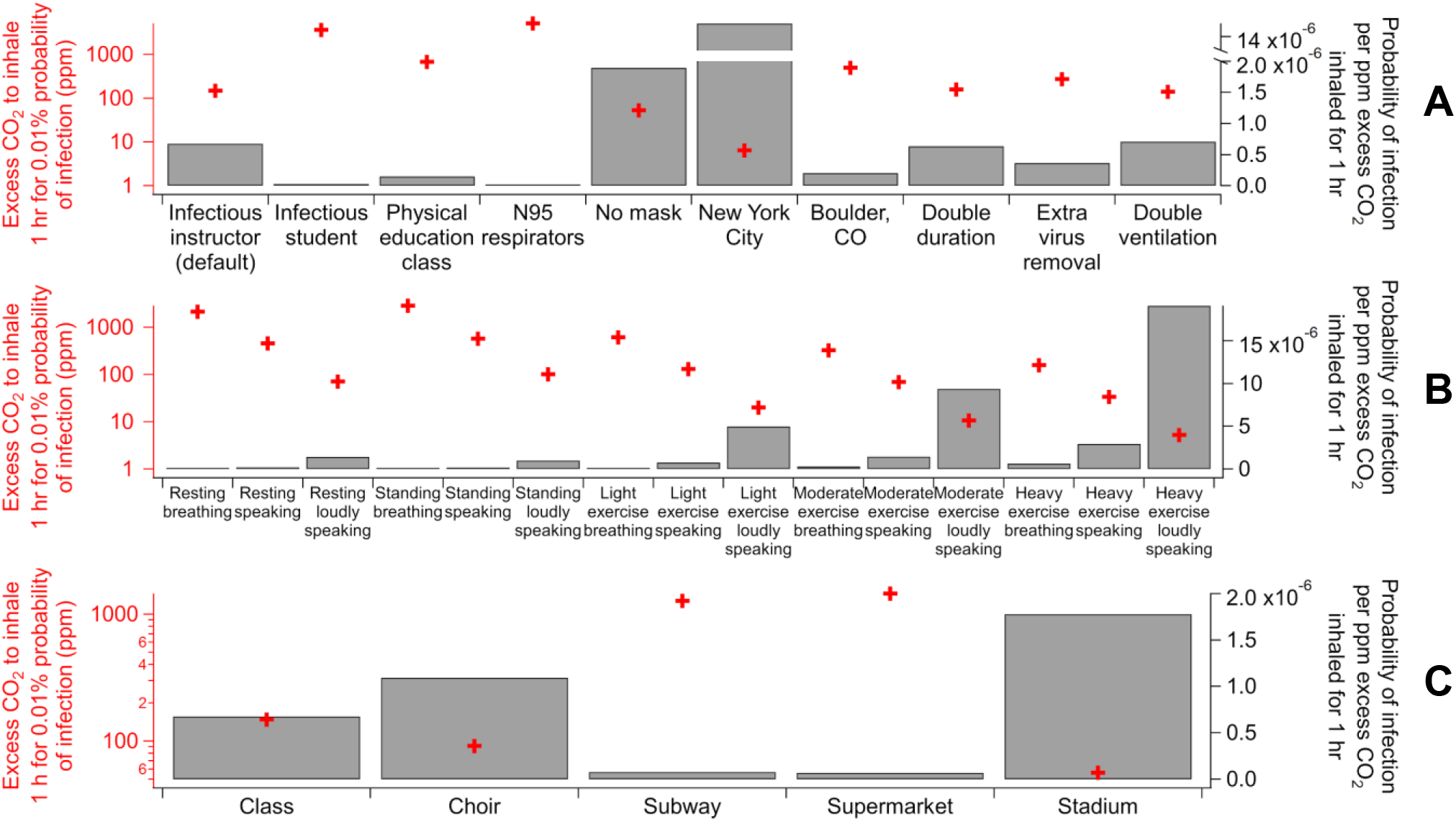
Excess CO_2_ volume mixing ratio (ppm) that an uninfected individual inhales for 1 h for a probability of infection of 0.01% 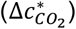 and probability of infection per ppm excess CO_2_ inhaled for 1 hr (inversely proportional to 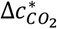) with a probability of an occupant being an infector of 0.1% (except the New York City and Boulder, CO cases in **A** and the Choir case in **C**) for (**A**) variants of the university class case (see Table S2 for the case details), (**B**) various activities (see Table S3 for details of the activities), and (**C**) several indoor environments (see Table S4 for the case details).

In the base class case, the infector is assumed to be the instructor. Compared to the case with a student being infector, 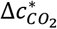 in the base case is ∼1.5 orders of magnitude lower, just because the vocalization of the instructor, who usually speaks, greatly enhances *E*_*p*_,^22,23^ while virus exhalation by students, who are assumed here to speak little, is much less efficient. In the case of a physical education (PE) class in the same indoor environment, where occupants are assumed to be doing heavy exercise and no talking, 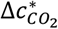 is much lower than for the infected student case in a traditional lecture (Fig. 2A). Compared to sitting, heavy exercise increases both occupants’ virus and CO_2_ exhalation rates to similar extents,^22–24^ which does not significantly change reference excess CO_2_ level. However, breathing rates of occupants doing intense activities are much higher than those sitting.^25^ Even if CO_2_ and SARS-CoV-2 concentrations are the same as in the infected student case, a susceptible person in the PE class case can still inhale a larger dose of SARS-CoV-2 and more excess CO_2_, and have a remarkably different *P*. As a result, a single recommendation of indoor CO_2_ threshold is not valid even for a series of school settings. The range of CO_2_ levels measured in real-world classrooms is very large.^26^ Reference excess CO_2_ level of the infectious student case (relatively safe) is exceeded in some classrooms, while that of the infectious instructor case (relatively risky) is met in other classrooms.

According to Eqs. 2 and 3, whether occupants wear masks and what masks they wear can make a substantial difference in infection risk through virus filtration in the same indoor setting. However, masks do not filter CO_2_. The base class case (with surgical masks), that with all occupants wearing N95 respirators, and that with no mask use have identical CO_2_ mixing ratios, but up to ∼2 orders of magnitude different probability of infection (Table S2) due to filtration of virus-containing particles by mask. Therefore, for the same probability of infection of 0.01%, the base class case is estimated by Eq. 7 to have a corresponding excess CO_2_ level x∼30 lower than the case with all occupants wearing N95 respirators, but x∼2 higher than the case with no mask use (Fig. 2A).

*η*_*I*_ is obviously another important factor governing the infection risk, as *P* proportional to it. Again, it has no impact on CO_2_. Compared to the base class case (*η*_*I*_ = 0.001), the estimated situations of similar classes in New York City (NYC) in April (*η*_*I*_ = 0.023) and in Boulder, CO in June (*η*_*I*_ = 0.0003) have x∼20 higher and x∼2 lower *P*, respectively (Table S2), and hence 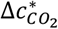 proportionally lower and higher, respectively (Fig. 2A). Note that 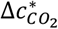 is smaller than the current typical accuracy of low-cost CO_2_ sensors (±50 ppm)^27^ and cannot be meaningfully measured by those sensors in very risky situations such as the NYC case here. Closure of environments with such low permissible 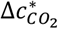 is likely needed. However, *η*_*im*_ usually cannot result in a difference in *P* greater than a factor of 2 under conditions of interest, since if *η*_*im*_ > 50%, the population has reached or is close to herd immunity^28^ and widespread transmission risk monitoring is no longer needed.

According to Eq. 7, the other variables that can affect 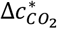 are *N, D, λ*, and *λ*_*0*_. 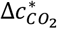 is generally not highly sensitive to them, although some of them (e.g., *λ*) can have a large impact on *P*. As long as occupants are not only a few, ^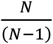^, where *N* plays a role in Eq. 7, is close to 1. The fraction term involving *D, λ*, and *λ*_*0*_ (after the product sign) in Eq. 7 usually does not deviate from 1 substantially (Fig. S2). It is close to 1 when *λD* is very small, and *λ*/*λ*_*0*_ when *λD* is very large. As long as the indoor environment is not very poorly ventilated nor equipped with very strong virus removal setups (e.g., substantial filtering of recirculated air, portable HEPA filters, germicidal UV), *λ*/*λ*_*0*_ is relatively close to 1. Compared to the base classroom case (*λ*/*λ*_*0*_ ∼ 1.3), doubling the duration or ventilation brings minimal changes to 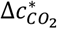. Increasing *λ*/*λ*_*0*_ to ∼3 by additional virus control measures increases 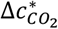 more significantly, as those measures do not remove CO_2_. But this change is still within a factor of 2 for the range of control measures in these examples (Fig. 2A).

As discussed above, occupants’ activities indoors, to which 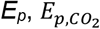, and *B* are all related, are a major or dominant factor governing the infection risk. We thus compile the data of these parameters as a function of activity (intensity and vocalization degree) (Table S3). Note that this compilation has large uncertainties from *E*_*p*_ data^22,23^ and matching of activity categories, which are all classified differently for 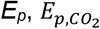, and *B* (see Section S3 for details). These uncertainties are currently difficult to quantify, but likely large enough to be the dominant uncertainty sources for the model output. Other sources of uncertainty are thus not discussed. Further systematic uncertainty analyses would be of interest. However, the trends shown are clear and thus able to reveal the *relative* risk of these activities with confidence. Simply, the stronger vocalization, the higher risk, and the more intense activity, the higher risk. We calculate 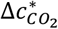 for these activities when *N* is large, *D* = 1 h, *η*_*I*_ = 0.001, *λ*_*0*_ = 3 h^-1^, *λ* = 4 h^-1^, no mask is used (Fig. 2B), a setting similar to the class case. Three class cases, i.e., base, infected student, and PE cases, can be easily related to the activity categories of “Standing – loudly speaking”, “Resting – breathing”, and “Heavy exercise – breathing”, respectively. The related pairs have 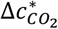 within x∼2 and their mask use setting and close but different 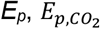, and *B* values can largely explain the differences in 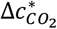.

Then we apply this analysis to a range of real-world settings, in addition to the class case, i.e., the Skagit County choir superspreading event,^5^ a subway car, a supermarket (focused on a worker), and an event in a stadium, which, though outdoors, often has somewhat stagnant air allowing virus-laden aerosols to build up and thus can be treated similarly as an indoor environment (see Table S4 for the specifications of these cases). Figures 2C and S1B shows their 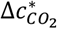 and 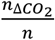, respectively. Again, these values span orders of magnitude. We can still relate these cases to the activity categories of “Standing – loudly speaking”, “Resting – breathing”, “Light exercise – breathing” (or “Light exercise – speaking”), and “Light exercise – speaking” (or “Light exercise – loudly speaking”), respectively.

For the actual choir case, its *η*_*I*_ is an order of magnitude lower than 0.1% while the estimated *E*_*p*_ is an order of magnitude higher (20), resulting in a similar reference excess CO_2_ level to that of “Standing – loudly speaking” shown in Fig. 2B. 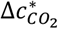 in the stadium case is between those of “Light exercise – speaking” and “Light exercise – loudly speaking”, as both activities may happen in the event. The difference of 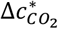 between the supermarket case and its related activities shown in Fig. 2B is mainly due to the long duration of the event (8 h). 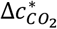 of the supermarket case *divided* by the duration leads to the excess CO_2_ threshold for the worker to inhale over 8 h between those of “Light exercise – breathing” and “Light exercise – speaking”. 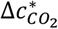 of the subway case is ∼1/3 lower than that of “Resting – breathing” in Fig. 2B because of the short duration (0.33 h) and mask use (universal use of surgical masks or equivalent).

As shown above, the infection risk analysis for various settings can be based on the relevant activities with adjustments for *η*_*I*_, *D*, mask use etc. For policy-making concerning acceptable indoor CO_2_ level, we also recommend an activity-dependent approach. Reference excess CO_2_ levels for indoor environments with certain types of activities mainly involved can be found in Fig. 2B. Then this mixing ratio can be scaled for typical *D* (by multiplying it) and target *P* (by multiplying its ratio to 0.01%) to obtain an excess CO_2_ threshold, which may be relaxed a little further depending on the local mask policy. The sum of this value and the local outdoor CO_2_ concentration, the latter of which we recommend measuring regularly due to possible variations,^29^ is the final recommended indoor CO_2_ concentration threshold. For more complex setups (e.g., with many CO_2_ meters in a company or school), a meter should be located outdoors to measure CO_2_ concentration continuously. To our knowledge, CO_2_ is the only quantity that can be easily measured by fast low-cost sensors as an infection risk proxy. The relative risk of infection in a given situation has been shown to scale with the excess CO_2_ concentration. The absolute risk can be estimated when the parameters needed are known. Calculations for various scenarios can be easily performed with the online COVID-19 aerosol transmission estimator.^30^ Then this method can provide a stronger scientific basis for using CO_2_, than having one threshold for all situations. However, it may still not be trivial for the general public to estimate the parameters used in our model and implement it. Regulatory authorities may derive the CO_2_ thresholds for different types of indoor spaces, or provide more assistance for businesses to do so. Even if the parameters are unknown, our study suggests that simply keeping the CO_2_ level and the physical intensity and vocalization level of the activities as low as practically feasible in indoor environments will still reduce the risk.

## Data Availability

All data is available in the main text or the supplementary materials. The COVID-19 Aerosol Transmission Estimator, with which a large part of calculations in this study were done, is freely available at https://tinyurl.com/covid-estimator.

## Acknowledgments

We thank Demetrios Pagonis and Bertrand Waucquez for useful discussions.

## References

(1) Organization, W. H. Coronavirus Disease (COVID-2019) Situation Reports; 2021.

(2) Prather, K. A., Marr, L. C., Schooley, R. T., McDiarmid, M. A., Wilson, M. E., Milton, D. K. Airborne Transmission of SARS-CoV-2. Science. 2020, 370 (6514), 303–304.

(3) Tang, J. W., Bahnfleth, W. P., Bluyssen, P. M., Buonanno, G., Jimenez, J. L., Kurnitski, J., Li, Y., Miller, S., Sekhar, C., Morawska, L., et al. Dismantling Myths on the Airborne Transmission of Severe Acute Respiratory Syndrome Coronavirus-2 (SARS-CoV-2). J. Hosp. Infect. 2021, 110, 89–96.

(4) Morawska, L., Milton, D. K. It Is Time to Address Airborne Transmission of Coronavirus Disease 2019 (COVID-19). Clin. Infect. Dis. 2020, in press.

(5) Miller, S. L., Nazaroff, W. W., Jimenez, J. L., Boerstra, A., Buonanno, G., Dancer, S. J., Kurnitski, J., Marr, L. C., Morawska, L., Noakes, C. Transmission of SARS-CoV-2 by Inhalation of Respiratory Aerosol in the Skagit Valley Chorale Superspreading Event. Indoor Air 2021, 31 (2), 314–323.

(6) Ma, J., Qi, X., Chen, H., Li, X., Zhang, Z., Wang, H., Sun, L., Zhang, L., Guo, J., Morawska, L., et al. Coronavirus Disease 2019 Patients in Earlier Stages Exhaled Millions of Severe Acute Respiratory Syndrome Coronavirus 2 Per Hour. Clin. Infect. Dis. 2020, in press.

(7) Liu, Y., Ning, Z., Chen, Y., Guo, M., Liu, Y., Gali, N. K., Sun, L., Duan, Y., Cai, J., Westerdahl, D., et al. Aerodynamic Analysis of SARS-CoV-2 in Two Wuhan Hospitals. Nature 2020, 582 (7813), 557–560.

(8) Lednicky, J. A., Lauzardo, M., Fan, Z. H., Jutla, A., Tilly, T. B., Gangwar, M., Usmani, M., Shankar, S. N., Mohamed, K., Eiguren-Fernandez, A., et al. Viable SARS-CoV-2 in the Air of a Hospital Room with COVID-19 Patients. Int. J. Infect. Dis. 2020, 100, 476–482.

(9) Bond, T. C., Bosco-Lauth, A., Farmer, D. K., Francisco, P. W., Pierce, J. R., Fedak, K. M., Ham, J. M., Jathar, S. H., VandeWoude, S. Quantifying Proximity, Confinement, and Interventions in Disease Outbreaks: A Decision Support Framework for Air-Transported Pathogens. Environ. Sci. Technol. 2021, in press.

(10) Ai, Z., Mak, C. M., Gao, N., Niu, J. Tracer Gas Is a Suitable Surrogate of Exhaled Droplet Nuclei for Studying Airborne Transmission in the Built Environment. Build. Simul. 2020, 13 (3), 489–496.

(11) Chen, W., Zhang, N., Wei, J., Yen, H.-L., Li, Y. Short-Range Airborne Route Dominates Exposure of Respiratory Infection during Close Contact. Build. Environ. 2020, 176 (January), 106859.

(12) Tellier, R., Li, Y., Cowling, B. J., Tang, J. W. Recognition of Aerosol Transmission of Infectious Agents: A Commentary. BMC Infect. Dis. 2019, 19 (1), 101.

(13) Qian, H., Miao, T., Liu, L., Zheng, X., Luo, D., Li, Y. Indoor Transmission of SARS-CoV-2. Indoor Air 2020, in press.

(14) Morawska, L., Tang, J. W., Bahnfleth, W., Bluyssen, P. M., Boerstra, A., Buonanno, G., Cao, J., Dancer, S., Floto, A., Franchimon, F., et al. How Can Airborne Transmission of COVID-19 Indoors Be Minimised? Environ. Int. 2020, 142, 105832.

(15) de Chaumont, F. On the Theory of Ventilation: An Attempt to Establish a Positive Basis for the Calculation of the Amount of Fresh Air Required for an Inhabited Air-Space. Proc. R. Soc. London 1875, 23, 187–201.

(16) Rudnick, S. N., Milton, D. K. Risk of Indoor Airborne Infection Transmission Estimated from Carbon Dioxide Concentration. Indoor Air 2003, 13 (3), 237–245.

(17) Martin, C. R., Zeng, N., Karion, A., Dickerson, R. R., Ren, X., Turpie, B. N., Weber, K. J. Evaluation and Environmental Correction of Ambient CO & Measurements from a Low-Cost NDIR Sensor. Atmos. Meas. Tech. 2017, 10 (7), 2383–2395.

(18) Mendell, M. J., Eliseeva, E. A., Davies, M. M., Spears, M., Lobscheid, A., Fisk, W. J., Apte, M. G. Association of Classroom Ventilation with Reduced Illness Absence: A Prospective Study in California Elementary Schools. Indoor Air 2013, 23 (6), 515–528.

(19) Jones, E., Young, A., Clevenger, K., Salimifard, P., Wu, E., Luna, M. L., Lahvis, M., Lang, J., Bliss, M., Azimi, P., et al. Healthy Schools: Risk Reduction Strategies for Reopening Schools; 2020.

(20) Cheng, S.-Y., Wang, C. J., Shen, A. C.-T., Chang, S.-C. How to Safely Reopen Colleges and Universities During COVID-19: Experiences From Taiwan. Ann. Intern. Med. 2020, M20–2927.

(21) Riley, E. C., Murphy, G., Riley, R. L. Airborne Spread of Measles in a Suburban Elementary School. Am. J. Epidemiol. 1978, 107 (5), 421–432.

(22) Buonanno, G., Stabile, L., Morawska, L. Estimation of Airborne Viral Emission: Quanta Emission Rate of SARS-CoV-2 for Infection Risk Assessment. Environ. Int. 2020, 141 (April), 105794.

(23) Buonanno, G., Morawska, L., Stabile, L. Quantitative Assessment of the Risk of Airborne Transmission of SARS-CoV-2 Infection: Prospective and Retrospective Applications. Environ. Int. 2020, 145, 106112.

(24) Persily, A., de Jonge, L. Carbon Dioxide Generation Rates for Building Occupants. Indoor Air 2017, 27 (5), 868–879.

(25) Chapter 6—Inhalation Rates. In Exposure Factors Handbook; US Environmental Protection Agency, 2011.

(26) Fromme, H., Heitmann, D., Dietrich, S., Schierl, R., Körner, W., Kiranoglu, M., Zapf, A., Twardella, D. Raumluftqualität in Schulen - Belastung von Klassenräumen Mit Kohlendioxid (CO 2), Flüchtigen Organischen Verbindungen (VOC), Aldehyden, Endotoxinen Und Katzenallergenen. Das Gesundheitswes. 2008, 70 (2), 88–97.

(27) Aranet. Wireless Indoor Air Monitoring, CO2, Temperature, Humidity Sensor https://aranet.com/product/aranet4-sensor/ (xaccessed Feb 26, 2021).

(28) Britton, T., Ball, F., Trapman, P. A Mathematical Model Reveals the Influence of Population Heterogeneity on Herd Immunity to SARS-CoV-2. Science. 2020, 369 (6505), 846–849.

(29) Dias Carrilho, J., Mateus, M., Batterman, S., Gameiro da Silva, M. Air Exchange Rates from Atmospheric CO2 Daily Cycle. Energy Build. 2015, 92, 188–194.

(30) Jimenez, J. L. COVID-19 Aerosol Transmission Estimator https://tinyurl.com/covid-estimator.

