## Supporting Information for "Exhaled CO_2_ as COVID-19 infection risk proxy for different indoor environments and activities"

**S1. Derivation of Eqs. 3 and 5 and limit values of the last fraction term in Eqs. 6 and 7**

The evolutions of both expected value of SARS-CoV-2 concentration ($\langle c \rangle$, in quanta m$^{-3}$) corresponding to a certain prevalence of infectors in local population ($\eta_I$) and excess $CO_2$ volume mixing ratio ($\Delta c_{CO_2}$) over time ($t$) can be expressed in terms of emission and loss rates:

$$\frac{d\langle c \rangle}{dt} = \frac{\langle E \rangle}{V} - \lambda \langle c \rangle \quad \text{(S1)}$$

$$\frac{d\Delta c_{CO_2}}{dt} = \frac{E_{CO_2}}{V} - \lambda_0 \Delta c_{CO_2} \quad \text{(S2)}$$

where $\langle E \rangle$ and $E_{CO_2}$ are the expected value of SARS-CoV-2 emission rate (in quanta h$^{-1}$) and excess $CO_2$ volume emission rate (m$^3$ h$^{-1}$), respectively. $\lambda \langle c \rangle$ and $\lambda_0 \Delta c_{CO_2}$ are the loss rates of SARS-CoV-2 and excess $CO_2$, respectively. $\lambda_0$ is the first-order of loss rate coefficient of excess $CO_2$ as $CO_2$ is only lost through ventilation. $\langle E \rangle$ and $E_{CO_2}$ can be further expanded as

$$\langle E \rangle = \eta_I (N-1) E_p (1 - m_{ex}) \quad \text{(S3)}$$

$$E_{CO_2} = N E_{p,CO_2} \quad \text{(S4)}$$

Equation S4 is easy to understand. $\eta_I(N-1)$ in Eq. S3 represents the expected value of the number of infectors. Since to calculate the probability of infection of a susceptible person, that person should be excluded from the occupants possibly being infectors, resulting in the $(N-1)$ term.

Inserting Eqs. S3 and S4 into Eqs. S1 and S2, respectively, and solving the resulted differential equations (under the assumption of no SARS-CoV-2 and of the same $CO_2$ concentration as outdoors initially) gives

$$\langle c \rangle = \frac{\eta_I (N-1) E_p (1-m_{ex})}{\lambda V} \left(1 - e^{-\lambda t}\right) \quad \text{(S5)}$$

$$\Delta c_{CO_2} = \frac{N E_{p,CO_2}}{\lambda_0 V} \left(1 - e^{-\lambda_0 t}\right) \quad \text{(S6)}$$

The averages of $\langle c \rangle$ and $\Delta c_{CO_2}$ during [0, $D$] are thus obtained below

$$\langle c_{avg} \rangle = \int_0^D \langle c \rangle dt = \frac{\eta_I (N-1) E_p (1-m_{ex})}{V} \cdot \left(\frac{1}{\lambda} - \frac{1-e^{-\lambda D}}{\lambda^2 D}\right) \quad \text{(S7)}$$

$$\Delta c_{avg,CO_2} = \int_0^D \Delta c_{CO_2} dt = \frac{N E_{p,CO_2}}{V} \cdot \left(\frac{1}{\lambda_0} - \frac{1-e^{-\lambda_0 D}}{\lambda_0^2 D}\right) \quad \text{(S8)}$$

When taking ratios between quantities related to $\langle c_{avg} \rangle$ and $\Delta c_{avg,CO_2}$, a large fraction term involving $\lambda$ and $\lambda_0$, $\left(\frac{1}{\lambda_0} - \frac{1-e^{-\lambda_0 D}}{\lambda_0^2 D}\right) / \left(\frac{1}{\lambda} - \frac{1-e^{-\lambda D}}{\lambda^2 D}\right)$, arises, as in Eqs. 6 and 7. This term approaches to 1 when $\lambda D$ is very small and $\lambda/\lambda_0$ when $\lambda D$ is very large. We show below the proof by applying L'Hôpital's rule repeatedly:

$$\lim_{\lambda D \to 0}\left[\left(\frac{1}{\lambda_0} - \frac{1-e^{-\lambda_0 D}}{\lambda_0^2 D}\right) / \left(\frac{1}{\lambda} - \frac{1-e^{-\lambda D}}{\lambda^2 D}\right)\right] = \lim_{\lambda D \to 0}\left[\left(\frac{\lambda_0}{\lambda}\right)^{-2}\left(\frac{\lambda_0}{\lambda}\lambda D - 1 + e^{-\frac{\lambda_0}{\lambda}\lambda D}\right) / \left(\lambda D - 1 + e^{-\lambda D}\right)\right]$$

$$= \lim_{\lambda D \to 0}\left[\left(\frac{\lambda_0}{\lambda}\right)^{-2}\left(\frac{\lambda_0}{\lambda} - \frac{\lambda_0}{\lambda}e^{-\frac{\lambda_0}{\lambda}\lambda D}\right) / \left(1 - e^{-\lambda D}\right)\right] = \lim_{\lambda D \to 0}\left[\left(\frac{\lambda_0}{\lambda}\right)^{-2}\left(\left(\frac{\lambda_0}{\lambda}\right)^{2} e^{-\frac{\lambda_0}{\lambda}\lambda D}\right) / e^{-\lambda D}\right]$$

$$= \lim_{\lambda D \to 0} e^{-\lambda D\left(1-\frac{\lambda_0}{\lambda}\right)} = e^0 = 1 \quad \text{(S9)}$$

Similarly,

$$\lim_{\lambda D \to \infty}\left[\left(\frac{1}{\lambda_0} - \frac{1-e^{-\lambda_0 D}}{\lambda_0^2 D}\right) / \left(\frac{1}{\lambda} - \frac{1-e^{-\lambda D}}{\lambda^2 D}\right)\right] = \lim_{\lambda D \to \infty}\left[\left(\frac{\lambda_0}{\lambda}\right)^{-2}\left(\frac{\lambda_0}{\lambda} - \frac{\lambda_0}{\lambda}e^{-\frac{\lambda_0}{\lambda}\lambda D}\right) / \left(1 - e^{-\lambda D}\right)\right]$$

$$= \lim_{\lambda D \to \infty}\left[\left(\frac{\lambda_0}{\lambda}\right)^{-1}\left(1 - e^{-\frac{\lambda_0}{\lambda}\lambda D}\right) / \left(1 - e^{-\lambda D}\right)\right] = \left(\frac{\lambda_0}{\lambda}\right)^{-1}(1-0)/(1-0) = \frac{\lambda}{\lambda_0} \quad \text{(S10)}$$

**S2. Volume mixing ratio of the excess $CO_2$ that an uninfected individual inhales for 1 h in that environment for certain infection risk**

Rudnick and Milton[1] derived the excess $CO_2$ concentration corresponding to $R_0$ of 1 ($\frac{E_{p,CO_2}}{E_p BD}$) for an aerosol-transmitted respiratory infectious disease during an indoor event under the assumptions of large $N$ and $\lambda \approx \lambda_0$. Unity $R_0$ is related to conditional probability of infection (for cases where one infector is present). $\eta_I$ is thus not considered in this type of problems. When Eq. 7 is applied under the assumptions of large $N$ and $\lambda \approx \lambda_0$, $\Delta c^*_{CO_2}$ is proportional to $\frac{E_{p,CO_2}}{E_p BD}$ (with $D$ = 1 h).

$\lambda \approx \lambda_0$ is a key approximation for convenient use of the Rudnick-Milton model, because this approximation allows the key quantity of this model, i.e., rebreathed fraction, to be considered identical for both virus-containing aerosols and $CO_2$. However, while trying to get to a more accurate and general model, we cannot start our analysis in this study based on this approximation. Therefore, the Rudnick-Milton model is not used in our derivation but discussed here.

**S3. $E_p$, $E_{p,CO2}$, and $B$ for different activities and associated uncertainties**

$E_p$, $E_{p,CO_2}$, and $B$ are all functions of activity according to the literature.[2–5] However, they have different domains in the literature studies. For $E_{p,CO_2}$, level of physical activity is quantified by a continuous variable, $M$ in MET (metabolic equivalent of task).[4] $E_{p,CO_2}$ is also a function of basic metabolic rate, which primarily depends on age, sex, body size, and body composition of a person.[4] For the data of $B$,[5] the levels of physical activity are discrete ("Sleep or Nap", "Sedentary/Passive", "Light Intensity", "Moderate Intensity", and "High Intensity"), and the data are also classified by age but not by sex.

To make the data of $E_{p,CO_2}$ and $B$ directly comparable, we take the averages of BMR for the males and females in the age ranges corresponding to the data of $B$ in ref 5, except for the range of 1–3 y as a single category (two categories, i.e., 1–2 and 2–3, in ref 5), and roughly assign "Sleep or Nap", "Sedentary/Passive", "Light Intensity", "Moderate Intensity", and "High Intensity" to $M$ = 1, 1.5, 2, 3.5, and 5 MET, respectively.[4] Then a quantity that involves $E_{p,CO_2}$ and $B$ and is critical for $\Delta c^*_{CO_2}$, $\frac{E_{p,CO_2}}{B}$, i.e., fraction of $CO_2$ in exhaled air, is calculated for the abovementioned discrete levels of physical activity for people in different age ranges (Fig. S3). At a specific physical activity level, $\frac{E_{p,CO_2}}{B}$ does not vary strongly with age for groups with age > 11 y (BMR > 6 MJ/d). Averages are thus taken for the groups with similar $\frac{E_{p,CO_2}}{B}$ at certain $M$ (Table S4). These averages are used in the infection risk analysis for different activities in the Main Text.

Buonanno et al.[2,3] estimated $E_p$ at different levels of physical activity as well as vocalization. In their estimates, there are only four levels of physical activities, i.e., "Resting", "Standing", "Light exercise", and "Heavy exercise". These four levels roughly correspond to "Sleep or Nap", "Sedentary/Passive", "Light Intensity", and "High Intensity". An interpolation is made by taking the geometric mean of $E_p$ at the "Light exercise", and "Heavy exercise" levels to generate data for $E_p$ at a "Moderate exercise" level, corresponding to "Moderate Intensity" for the $B$ data (Table S4). The dimension of vocalization for the $E_p$ data from Buonanno et al. is preserved in this study, as degree of vocalization is critical in determining $E_p$.[2,3] For all activities listed in Fig. 2*B* and Table S4, the data of $E_p$ and $\frac{E_{p,CO_2}}{B}$ are now available and the relevant infection risk analysis can be done.

Large uncertainties are associated with the data of $E_p$ and $\frac{E_{p,CO_2}}{B}$ in Table S4. The $E_p$ estimates themselves are highly uncertain, with possible ranges often spanning over an order of magnitude.[2,3] The discretization of physical activity level can also be a major uncertainty source, as there are only five discrete levels to cover the domain of a continuous variable $M$.

**References:**

(1) Rudnick, S. N.; Milton, D. K. Risk of Indoor Airborne Infection Transmission Estimated from Carbon Dioxide Concentration. *Indoor Air* **2003**, *13* (3), 237–245.

(2) Buonanno, G.; Stabile, L.; Morawska, L. Estimation of Airborne Viral Emission: Quanta Emission Rate of SARS-CoV-2 for Infection Risk Assessment. *Environ. Int.* **2020**, *141* (April), 105794.

(3) Buonanno, G.; Morawska, L.; Stabile, L. Quantitative Assessment of the Risk of Airborne Transmission of SARS-CoV-2 Infection: Prospective and Retrospective Applications. *Environ. Int.* **2020**, *145*, 106112.

(4) Persily, A.; de Jonge, L. Carbon Dioxide Generation Rates for Building Occupants. *Indoor Air* **2017**, *27* (5), 868–879.

(5) Chapter 6—Inhalation Rates. In *Exposure Factors Handbook*; US Environmental Protection Agency, 2011.
(6) Davies, A.; Thompson, K.-A.; Giri, K.; Kafatos, G.; Walker, J.; Bennett, A. Testing the Efficacy of Homemade Masks: Would They Protect in an Influenza Pandemic? *Disaster Med. Public Health Prep.* **2013**, *7* (4), 413–418.
(7) Bhangar, S.; Huffman, J. A.; Nazaroff, W. W. Size- resolved Fluorescent Biological Aerosol Particle Concentrations and Occupant Emissions in a University Classroom. *Indoor Air* **2014**, *24* (6), 604–617.
(8) van Doremalen, N.; Bushmaker, T.; Morris, D. H.; Holbrook, M. G.; Gamble, A.; Williamson, B. N.; Tamin, A.; Harcourt, J. L.; Thornburg, N. J.; Gerber, S. I.; et al. Aerosol and Surface Stability of SARS-CoV-2 as Compared with SARS-CoV-1. *N. Engl. J. Med.* **2020**, *382* (16), 1564–1567.
(9) Thatcher, T. L.; Lai, A. C. K.; Moreno-Jackson, R.; Sextro, R. G.; Nazaroff, W. W. Effects of Room Furnishings and Air Speed on Particle Deposition Rates Indoors. *Atmos. Environ.* **2002**, *36* (11), 1811–1819.
(10) Miller, S. L.; Nazaroff, W. W.; Jimenez, J. L.; Boerstra, A.; Buonanno, G.; Dancer, S. J.; Kurnitski, J.; Marr, L. C.; Morawska, L.; Noakes, C. Transmission of SARS- CoV- 2 by Inhalation of Respiratory Aerosol in the Skagit Valley Chorale Superspreading Event. *Indoor Air* **2021**, *31* (2), 314–323.
(11) Veres, P. R.; Faber, P.; Drewnick, F.; Lelieveld, J.; Williams, J. Anthropogenic Sources of VOC in a Football Stadium: Assessing Human Emissions in the Atmosphere. *Atmos. Environ.* **2013**, *77*, 1052–1059.

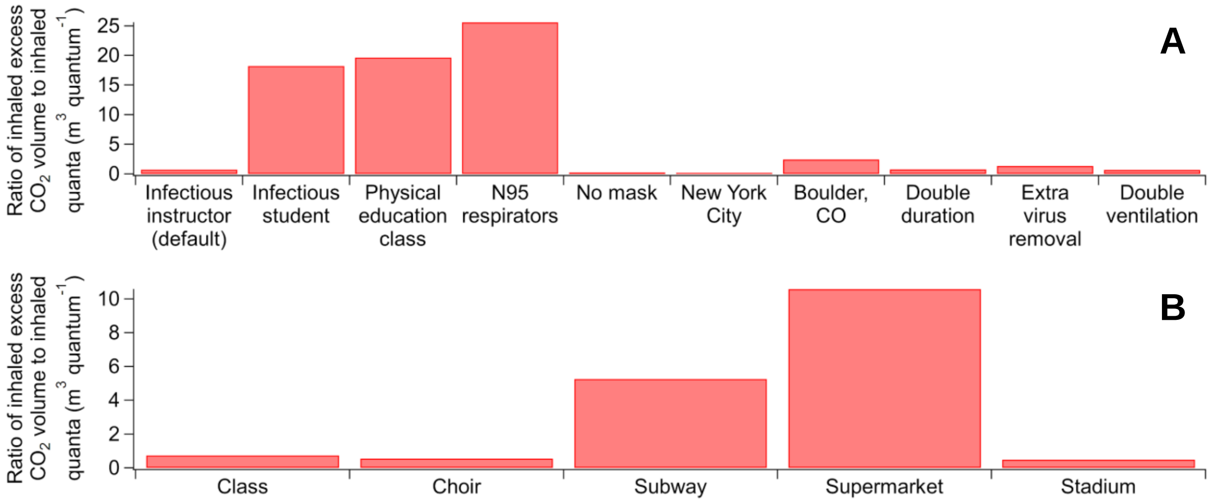


**Figure S1.** Ratio of inhaled excess $CO_2$ volume to inhaled SARS-CoV-2 quanta ($m^3$ $quantum^{-1}$) for (A) variants of the university class case (see Table S2 for the case details) and (B) several indoor environments (see Table S4 for the case details).

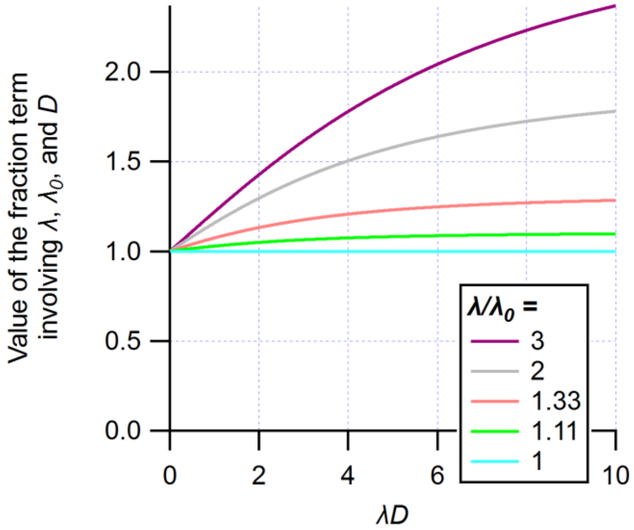


**Figure S2.** Value of the fraction term involving $\lambda$, $\lambda_0$, and $D$ in Eqs. 6 and 7 as a function of $\lambda D$.

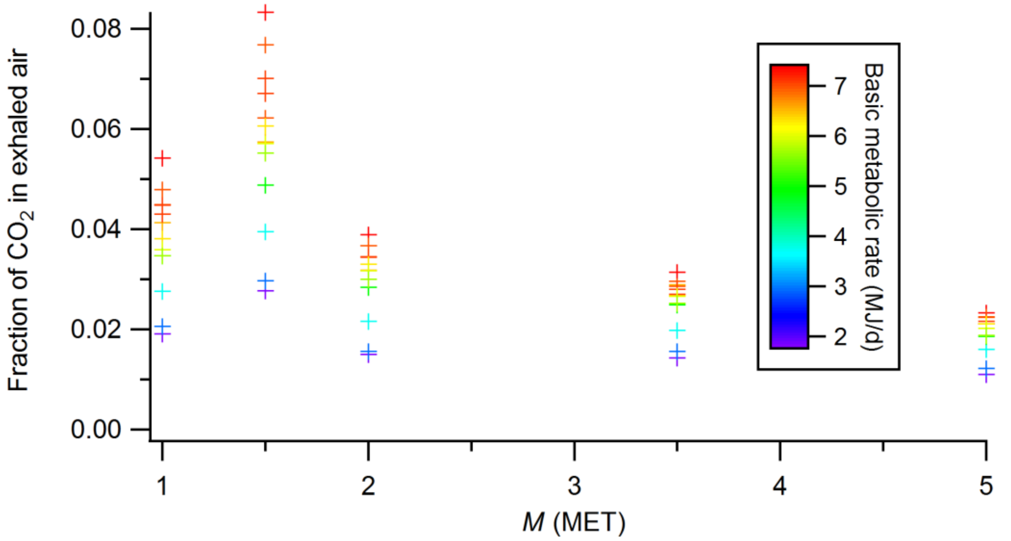


**Figure S3.** Fraction of $CO_2$ in exhaled air at several physical activity levels (represented by the variable *M*; see Materials and Methods for detail) for different age groups (colored by corresponding basic metabolic rates).

**Table S1.** Symbols used in this study.

| *Symbol* | *Physical meaning* | *Unit (dimension -less if no unit indicated)* |
|---|---|---|
| $B$ | Breathing rate of the susceptible person | $m^3\ h^{-1}$ |
| $c_{avg}$ | Average virus concentration in the air over the duration of the event | quanta $m^{-3}$ |
| $\langle c_{avg} \rangle$ | Expected value of $c_{avg}$, when an occupant has a probability of being immune | quanta $m^{-3}$ |
| $\Delta c_{avg,CO_2}$ | Average excess $CO_2$ volume mixing ratio | |
| $\Delta c^*_{CO_2}$ | Volume mixing ratio of the excess $CO_2$ that an uninfected individual inhales for 1 h in an environment with $\eta_I$ = 0.1% for $P$ = 0.01% | |
| $D$ | Duration of the event | h |
| $E_p$ | SARS-CoV-2 exhalation rate by an infector | quanta $h^{-1}$ |
| $E_{p,CO_2}$ | $CO_2$ exhalation rate per person | $m^3\ h^{-1}$ |
| $\eta_I$ | Probability of an occupant being an infector | |
| $\eta_{im}$ | Probability of an occupant being immune | |
| $\lambda$ | First-order overall rate constant of the virus infectivity loss | $h^{-1}$ |
| $\lambda_0$ | Ventilation rate | $h^{-1}$ |
| $m_{ex}$ | Mask filtration efficiency for exhalation | |
| $m_{in}$ | Mask filtration efficiency for inhalation | |
| $N$ | Number of occupants | |
| $n$ | Amount of the virus infectious doses (“quanta”) inhaled by a susceptible person in a given indoor environment | quanta |
| $\langle n \rangle$ | Expected value of $n$, when an occupant has a probability of being immune | quanta |
| $n_{\Delta CO_2}$ | Inhaled excess (human-exhaled) $CO_2$ volume | $m^3$ |
| $P$ | Probability of infection of a susceptible person | |
| $V$ | Indoor environment volume | $m^3$ |
| *(Below are the symbols that appear in the SI only)* | | |
| $\langle c \rangle$ | Expected value of virus concentration, when an occupant has a probability of being immune | quanta $m^{-3}$ |
| $\Delta c_{CO_2}$ | Excess $CO_2$ volume mixing ratio | |

| | | |
|---|---|---|
| $\langle E \rangle$ | Expected value of virus emission rate | quanta $h^{-1}$ |
| $E_{CO_2}$ | Excess $CO_2$ volume emission rate | $m^3$ $h^{-1}$ |
| $t$ | Time | h |

**Table S2.** Input parameter settings (number of occupants, $N$; volume of the indoor environment, $V$; SARS-CoV-2 exhalation rate, $E_p$; breathing rate, $B$; probability of an occupant being immune, $\eta_{im}$; probability of an occupant being infector, $\eta_I$; duration, $D$; mask filtration efficiency for exhalation, $m_{ex}$; mask filtration efficiency for inhalation, $m_{in}$; ventilation rate, $\lambda_0$; first-order SARS-CoV-2 loss rate coefficient, $\lambda$) of the case for a typical university class and of its variations. Model results for the expected value of the amount of SARS-CoV-2 inhaled by an uninfected individual ($\langle \boldsymbol{n} \rangle$), average excess $CO_2$ volume mixing ratio ($\Delta \boldsymbol{c}_{\boldsymbol{avg,CO_2}}$), and the volume mixing ratio of the excess $CO_2$ that an uninfected individual inhales for 1 h in that environment for a probability of infection of 0.01% ($\Delta \boldsymbol{c}^*_{\boldsymbol{CO_2}}$) of these cases are also shown. See footnotes for details on the estimation of some parameter values.

| Case | $N$ | $V$ ($m^3$) | $E_p$ (quanta $h^{-1}$) | $E_{p,CO_2}$ ($m^3$ $h^{-1}$) | $B$ ($m^3$ $h^{-1}$) | $\eta_{im}$ | $\eta_I$ | $D$ (h) | $m_{ex}$ | $m_{in}$ | $\lambda_0$ ($h^{-1}$) | $\lambda$ ($h^{-1}$) | $\langle n \rangle$ (quanta) | $\Delta c_{avg,CO_2}$ (ppm) | $\Delta c^*_{CO_2}$ (ppm) |
|---|---|---|---|---|---|---|---|---|---|---|---|---|---|---|---|
| Infectious instructor (default) | 10 | 142 | 100 | 0.0203 | 0.516 | 0 | 0.001 | 0.833 | 0.5 | 0.3 | 3 | 3.92 | 1.70E-04 | 302 | 148 |
| Infectious student | 10 | 142 | 4 | 0.0203 | 0.516 | 0 | 0.001 | 0.833 | 0.5 | 0.3 | 3 | 3.92 | 6.87E-06 | 302 | 3670 |
| Physical education class | 10 | 142 | 13.5 | 0.0732 | 3 | 0 | 0.001 | 0.833 | 0.5 | 0.3 | 3 | 3.92 | 1.34E-04 | 1089 | 678 |
| N95 respirators | 10 | 142 | 100 | 0.0203 | 0.516 | 0 | 0.001 | 0.833 | 0.9 | 0.9 | 3 | 3.92 | 4.91E-06 | 302 | 5130 |
| No mask | 10 | 142 | 100 | 0.0203 | 0.516 | 0 | 0.001 | 0.833 | 0 | 0 | 3 | 3.92 | 4.78E-04 | 302 | 52.7 |
| New York City | 10 | 142 | 100 | 0.0203 | 0.516 | 0 | 0.023 | 0.833 | 0.5 | 0.3 | 3 | 3.92 | 3.91E-03 | 302 | 6.45 |
| Boulder, CO | 10 | 142 | 100 | 0.0203 | 0.516 | 0 | 0.0003 | 0.833 | 0.5 | 0.3 | 3 | 3.92 | 5.11E-05 | 302 | 493 |
| Double duration | 10 | 142 | 100 | 0.0203 | 0.516 | 0 | 0.001 | 1.667 | 0.5 | 0.3 | 3 | 3.92 | 4.03E-04 | 383 | 158 |
| Extra virus removal | 10 | 142 | 100 | 0.0203 | 0.516 | 0 | 0.001 | 0.833 | 0.5 | 0.3 | 3 | 8.92 | 9.22E-05 | 302 | 273 |
| Double ventilation | 10 | 142 | 100 | 0.0203 | 0.516 | 0 | 0.001 | 0.833 | 0.5 | 0.3 | 6 | 6.92 | 1.13E-04 | 191 | 141 |

$E_p$, $E_{p,CO_2}$, and $B$: see Section S3.

$\eta_I$: the values for New York City at the peak of the first COVID-19 wave in spring 2020, and the value for Boulder, CO during a period of low disease prevalence in summer 2020 are estimated based on the New York Times Coronavirus Database (https://www.nytimes.com/article/coronavirus-county-data-us.html). A typical value in between is assumed for all other cases.

$m_{ex}$ and $m_{in}$: mask parameters are estimated based on Davies et al.[6] $m_{in}$ is assigned a value lower than reported by Davies et al.[6] given imperfect wearing and fit in the community.

$\lambda_0$: a typical value is chosen within the range reported by Bhangar et al.[7]

$\lambda$: see above for ventilation ($\lambda_0$). Removal rates due to virus infectivity decay and aerosol deposition are estimated based on van Doremalen et al. [8] and Thatcher et al.,[9] respectively.

**Table S3.** SARS-CoV-2 exhalation rate ($E_p$), fraction of $CO_2$ in exhaled air ($\frac{E_{p,CO_2}}{B}$), and volume mixing ratio of the excess $CO_2$ that an uninfected individual inhales for 1 h in that environment for a probability of infection of 0.01% ($\Delta c^*_{CO_2}$) for activities with different physical and vocal levels. $\Delta c^*_{CO_2}$ is estimated with probability of an occupant being infector of 0.1% and ventilation accounting for all SARS-CoV-2 loss.

| Activity | $E_p$ (quanta $h^{-1}$) | $\frac{E_{p,CO_2}}{B}$ | $\Delta c^*_{CO_2}$ (ppm) |
|---|---|---|---|
| Resting – breathing | 2 | 0.0428 | 2140 |
| Resting – speaking | 9.4 | 0.0428 | 455 |
| Resting – loudly speaking | 60.5 | 0.0428 | 70.7 |
| Standing – breathing | 2.3 | 0.0656 | 2850 |
| Standing – speaking | 11.4 | 0.0656 | 575 |
| Standing – loudly speaking | 65.1 | 0.0656 | 101 |
| Light exercise – breathing | 5.6 | 0.0342 | 611 |
| Light exercise – speaking | 26.3 | 0.0342 | 130 |
| Light exercise – loudly speaking | 170 | 0.0342 | 20.1 |
| Moderate exercise – breathing | 8.7 | 0.0280 | 322 |
| Moderate exercise – speaking | 40.7 | 0.0280 | 68.8 |
| Moderate exercise – loudly speaking | 263 | 0.0280 | 10.7 |
| Heavy exercise – breathing | 13.5 | 0.0214 | 158 |
| Heavy exercise – speaking | 63.1 | 0.0214 | 33.8 |
| Heavy exercise – loudly speaking | 408 | 0.0214 | 5.23 |

**Table S4.** Same format as Table S2, but for different environments, i.e., a university class, the Skagit County choir superspreading event, a subway car, a supermarket (focused on a worker), and an event in a stadium. Values of parameters are typical of real environments that we have analyzed (see footnotes for detail).

| Case | $N$ | $V$ (m$^3$) | $E_p$ (quanta h$^{-1}$) | $E_{p,CO_2}$ (m$^3$ h$^{-1}$) | $B$ (m$^3$ h$^{-1}$) | $\eta_{im}$ | $\eta_I$ | $D$ (h) | $m_{ex}$ | $m_{in}$ | $\lambda_0$ (h$^{-1}$) | $\lambda$ (h$^{-1}$) | $\langle n \rangle$ (quanta) | $\Delta c_{avg,CO_2}$ (ppm) | $\Delta c^*_{CO_2}$ (ppm) |
|---|---|---|---|---|---|---|---|---|---|---|---|---|---|---|---|
| Class | 10 | 142 | 100 | 0.0203 | 0.516 | 0 | 0.001 | 0.833 | 0.5 | 0.3 | 3 | 3.92 | 1.70E-04 | 302 | 148 |
| Choir | 61 | 810 | 970 | 0.0370 | 1.56 | 0 | 0.00011 | 2.5 | 0 | 0 | 0.7 | 1.62 | 1.42E-02 | 2102 | 91.3 |
| Subway | 35 | 150 | 25 | 0.0285 | 0.42 | 0.15 | 0.001 | 0.333 | 0.5 | 0.3 | 5.7 | 10.22 | 1.66E-05 | 645 | 1270 |
| Super-market | 75 | 2040 | 10 | 0.0275 | 0.72 | 0 | 0.001 | 8 | 0.5 | 0.3 | 3 | 3.92 | 1.69E-04 | 322 | 1450 |
| Stadium | 31000 | 255000 | 50 | 0.0248 | 0.72 | 0 | 0.001 | 1.5 | 0 | 0 | 40 | 40.92 | 1.58E-04 | 74 | 56.2 |

The class case represents a university classroom at the University of Colorado Boulder in Fall 2020 with COVID-19 mitigation measures. The choir case is the rehearsal that led to the outbreak at Skagit county in March 2020.[10] The subway case uses real data provided by a public transport operator in a large North American City (not identified due to confidentiality). The supermarket is located in Colorado (not identified due to confidentiality). The stadium parameters are estimated from the study of Veres et al.[11]
